# National scale-up of etiological testing for *N. gonorrhoeae* and *C. trachomatis* in South Africa: a health economic modelling analysis

**DOI:** 10.1101/2025.10.10.25337253

**Authors:** Joshua M Chevalier, Sarah J Girdwood, Megan A Hansen, Lise Jamieson, Nkgomeleng Lekodeba, Katherine Snyman, Regina Maithufi, Birgitta Gleeson, Benjamin Blumel, Shaukat Khan, Kyra H Grantz, Brooke E Nichols

## Abstract

**Background:** Neisseria gonorrhoeae (NG) and Chlamydia trachomatis (CT) remain highly prevalent in South Africa, where syndromic management is the standard-of-care (SOC) for sexually transmitted infections (STIs). However, syndromic management lacks diagnostic precision leading to both under-treatment and excess antibiotic use, which contributes to antimicrobial resistance (AMR). With a novel lateral flow assay (LFA) for NG approaching market entry and an extensive existing GeneXpert network in South Africa, there is an opportunity to shift towards etiology-informed management of STIs.

**Methods:** We developed a static cohort model to evaluate the health and economic outcomes of scaling up testing for NG/CT across symptomatic and opportunistic screening pathways in South Africa. Scenarios included syndromic management, GeneXpert testing, and hypothetical NG/CT point-of-care tests (POCTs). Outcomes assessed included cases treated, quality-adjusted life years (QALYs) gained, excess antibiotic use, and cost-effectiveness.

**Results:** Syndromic management produced the poorest health outcomes and highest antibiotic overuse. POCTs achieved the best health outcomes and were cost-effective compared to SOC, with an incremental cost-effectiveness ratio (CER) of US$1,083–$3,497 per QALY gained. GeneXpert, though dominated overall, could be considered cost-effective for testing of symptomatic women (Average CER: US$5,073). Diagnostic testing had the greatest benefit among women due to poor sensitivity of syndromic management. Opportunistic screening using a NG-only POCT during antenatal care, family planning clinics or HIV-related services was also found to be cost-effective.

**Conclusion:** Despite higher costs, etiological testing offers significant benefits over syndromic management— particularly for women—by improving diagnostic accuracy, reducing unnecessary antibiotic use, and supporting antibiotic stewardship. As South Africa considers introducing new treatments like zoliflodacin, investment in diagnostic testing is essential to preserve treatment efficacy and reduce long-term STI burden.

## Introduction

Curable bacterial sexually transmitted infections (STIs) remain a significant global health burden. The World Health Organization (WHO) estimated 211 million new *Neisseria gonorrhoeae* (NG) and *Chlamydia trachomatis* (CT) infections in 2020 [1]. This burden falls disproportionately on low- and middle-income countries, where a lack of laboratory resources and infrastructure make etiological diagnosis and treatment challenging. South Africa experiences some of the highest rates of NG/CT infections in the African region, where prevalence of NG and CT were estimated at 6.6% and 14.7% among women, and 3.5% and 6.0% among men, respectively [2,3].

In South Africa, of those with STI-related symptoms, 64% of men and 70% of women seek treatment through the public sector where STIs are diagnosed and treated through syndromic management [4]. While current STI guidelines indicate treatment for all individuals with urethral or vaginal discharge syndromes (UDS/VDS), studies have shown low sensitivity and specificity of syndromic management, particularly among women [5,6]. This is compounded by the fact many women experiencing VDS do not have NG/CT, resulting in antibiotic overuse and contributing to antimicrobial resistance (AMR) [7]. The WHO categorizes NG as a high-priority antibiotic-resistant pathogen, as resistance to ceftriaxone—the current last line empiric treatment option— has been documented [8]. Zoliflodacin and gepotidacin, potential new antibiotics to treat NG, show high activity against the pathogen, including resistant isolates [9,10]. These antibiotics are expected to become available in the coming years, and their efficacy will need to be protected through better diagnostics and stringent use [11,12].

A significant limitation of syndromic management is its inability to detect the high proportion of asymptomatic cases in both men and women [13,14]. Consequently, most cases will remain undetected in the absence of asymptomatic screening, sustaining transmission. Untreated NG/CT can reduce quality of life, contribute to HIV transmission, lead to the development of pelvic inflammatory disease (PID) and infertility in women, or result in pregnancy-related complications such as vertical transmission and preterm birth [15]. Key populations could be considered for opportunistic screening based on their vulnerability, health consequences, or attributable disease and transmission burden; these include adolescent girls and young women (AGYW), pregnant women, female sex workers (FSW), men who have sex with men (MSM), transgender women (TGW), and people living with HIV (PLHIV) [16].

The current gold standard diagnostic for gonorrhea and chlamydia is polymerase chain reaction (PCR) through molecular assays such as GeneXpert, which has a high reported sensitivity and specificity [17]. The high cost of these assays, combined with delays to results—preventing immediate treatment—generally makes implementation less feasible in limited-resource settings. However, South Africa has a large network of GeneXpert devices used for tuberculosis testing that could be utilized for diagnosis of NG/CT. Alternatively, rapid point-of-care tests (POCTs) that abide by the ASSURED criteria (Affordable, Sensitive, Specific, User-friendly, Rapid and Robust, Equipment-free, and Deliverable) could replace syndromic management to accurately treat infections, reduce antibiotic overuse, and improve health outcomes, while maintaining cost-effectiveness [18]. Early evaluations of a novel lateral flow POCT for NG show promising sensitivity and specificity [19]. Here, we present a modelling and health economic evaluation of the national scale up of symptomatic and asymptomatic testing for NG/CT using a novel POCT or GeneXpert test in South Africa.

## Methods

We modelled NG/CT diagnosis and treatment for a static cohort representing the national South African population who would be expected to seek care for STI-related symptoms in the public sector over a one-year period. Diagnosis occurred through syndromic management without diagnostic testing, a novel POCT for NG-only or both NG/CT, and GeneXpert for NG/CT. We then modelled opportunistic screening of target populations through five different use cases using POCT or GeneXpert. Costs of STI care-seeking and treatment were estimated from the provider perspective and, together with modelled health outcomes, were used to calculate the incremental cost-effectiveness ratios per additional quality adjusted life-year (QALY) gained to compare diagnostic strategies. The model and analysis were performed in Microsoft Excel (Version: 16.90.2), while figures were generated using R 4.4.2. This study follows the Consolidated Health Economic Evaluation Reporting Standards (CHEERS; **Table S1**).

### Population

We modeled the national population of men and women aged 15-59 in South Africa (N= 37,993,000), stratified by key populations—AGYW aged 15-24, women aged 25-59, FSW, pregnant women, MSM, men who have sex with women (MSW) aged 15-59, TGW, and PLHIV. The population breakdown by group—inclusive of overlap between populations—is shown in **Table 1**. All data used in the parameterization of the modelled population was publicly available.

**Table 1.**
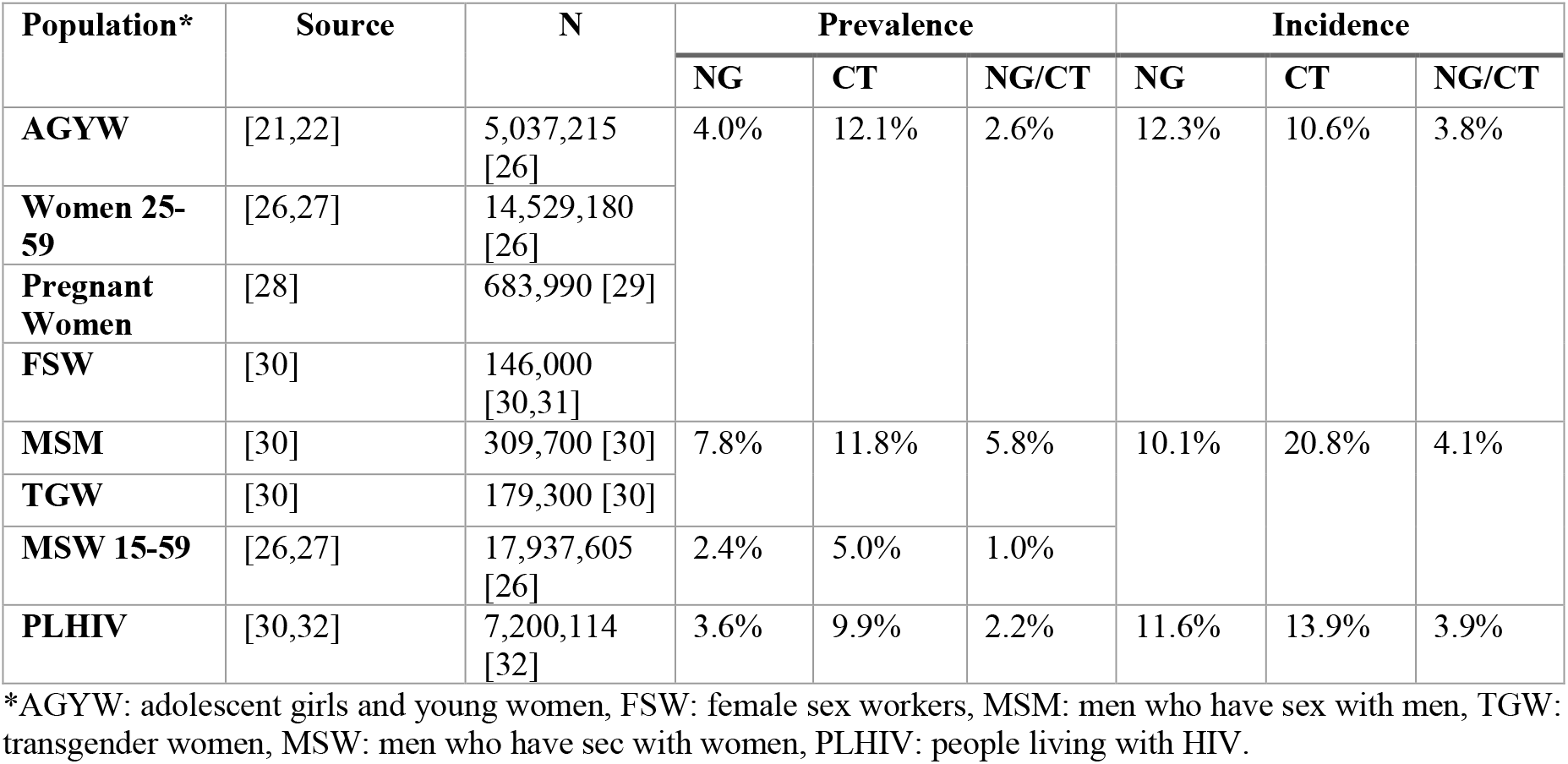
Target population size (inclusive of overlap between groups), NG, CT, and NG/CT co-infection prevalence and incidence.

### Gonorrhea and chlamydia prevalence, incidence, and care-seeking

NG/CT incidence and prevalence in South Africa were defined by key population when evidence was available, otherwise broader population estimates were used (**Table 1**) [2,20,21]. NG/CT incidence and prevalence were adjusted to include co-infection [22]. The symptomatic proportion of NG was 65% among men and 25% among women, while the symptomatic proportion of CT was 22% and 12%, respectively [23]. The symptomatic proportion of NG/CT represented 93% of all men presenting with UDS and 29% of all women with VDS [7,24].

NG/CT annual cumulative incidence was used to determine the number of symptomatic individuals who sought STI testing and treatment. The total number of symptomatic individuals was inflated using the relative proportions of UDS/VDS with NG/CT etiology among men and women, respectively. We assumed 64% of men and 70% of women sought care for their STI symptoms, and 70% of those did so through the public sector [4]. We assumed individuals with true NG/CT infection, who received a false negative diagnosis, returned for a follow-up visit with lost-to-follow-up (LTFU) rates of 71% among men and 51% among women [25].

### Syndromic Management

Syndromic management is the standard-of-care (SOC) for STIs in South Africa. All symptomatic individuals presenting to the public sector with UDS and VDS undergo a clinical evaluation that includes a speculum examination for female patients. Syndromic management consists of empiric treatment with ceftriaxone and doxycycline for NG/CT [16]. Syndromic management is associated with a sensitivity of 91.5% and specificity of 60.3% among men, and 44.9% and 74.2% among women, respectively [6,33]. Syndromic management sensitivity and specificity were modified in a sensitivity analysis (**Text S1**).

### Diagnostic Tests

A novel NG lateral flow assay (LFA) that generates results within 30 minutes was evaluated at the primary healthcare level in South Africa, revealing a sensitivity of 96.1% and specificity of 97.2% from urine samples [19]. In this analysis, we assume a comparable LFA for both NG and CT with the same sensitivity and specificity for the POCT. Additionally, we evaluated the impact of the NG-only LFA, where symptomatic individuals who test NG-negative (CT-suspect cases) are treated under the SOC for CT infection. Sensitivity and specificity were reduced in a sensitivity analysis (**Text S1**).

GeneXpert testing was assumed to be conducted in central laboratories in accordance with South African GeneXpert infrastructure for tuberculosis testing [34]. GeneXpert sensitivity and specificity were assumed to be 95% and 100% for NG, and 94% and 99% for CT, respectively [17]. GeneXpert results were expected to be returned no sooner than one day following testing, creating potential for LTFU. We assumed 8% of individuals would be LTFU and not receive the appropriate diagnosis and treatment [35]. LTFU was varied in a sensitivity analysis (**Text S1**).

### Opportunistic Screening Scenarios

NG/CT prevalence was used to determine the number of symptomatic and asymptomatic individuals who may be captured passively through opportunistic screening strategies with the NG-only POCT. In these opportunistic screening scenarios, prevalence was used in place of incidence as we assume the difference between prevalence and incidence represents the proportion of NG/CT cases that were captured and treated through symptomatic testing. We identified five use-cases (settings where STI screening could be feasibly implemented) for opportunistic screening—antenatal care (ANC), family planning, pre-exposure prophylaxis (PrEP) initiation, HIV testing, and HIV care. The estimated proportion of each target population that utilizes each use-case, as well as the proportions that seek care through the public sector in each use-case are outlined in **Supplementary Table S2**. As these use-cases assume passive screening, in the NG-only POCT scenarios, we do not assume any syndromic management or CT treatment for symptomatic NG-negative individuals.

### Health Outcomes

The primary outcome in this analysis was the number of NG/CT cases correctly treated. Secondary outcomes included excess antibiotics use (for those that received antibiotics without an underlying infection), PID, vertical transmission events (ophthalmia neonatorum and pneumonia), and preterm births. The various health-states—asymptomatic infection, symptomatic infection, PID, vertical transmission events, and pre-term births—were assigned utility weights, and total QALYs per scenario were calculated (**Supplementary Tables S3, S4**). The QALY values represented the average QALY associated with the respective health-state, and total QALYs were calculated over a time horizon of one year. QALY estimates were not specific to South Africa but were derived from publicly available data. QALYs were used in lieu of disability-adjusted life years (DALYs) due to data availability on the utility scores for the specific health-states used in this analysis— vertical transmission events, preterm birth, PID, and symptomatic versus asymptomatic infection.

### Costs

We conducted an economic cost analysis from the provider perspective, representing the South African National Department of Health (**Table 2**). Each client was assumed to have one visit covering triage, screening/diagnosis, and treatment, unless GeneXpert testing was conducted, in which case an additional visit was required for result delivery and treatment initiation. NG and NG/CT POCT test costs included a professional nurse’s time, training, and the cost of the test, based on the minimal and optimal requirements in the WHO target product profile (TPP) for STI POCTs [36]. The minimal price of $12 was used for a NG/CT POCT and the optimal price of $3 for the NG-only POCT. GeneXpert diagnostic costs included sample registration, equipment, human resources, supplies, laboratory overheads, and external quality assessment. Staff, equipment, and consumables were based on a recent GeneXpert MTB/XDR costing analysis in a high-throughput lab in Gauteng [37]. GeneXpert cartridge and swab costs for NG/CT were obtained from the manufacturer [38]. Equipment costs were annualized over five years with a 4% discount rate [39]. Treatment regimens were based on the latest WHO STI guidelines (July 2024) with drug prices sourced from South African tenders [40,41]. Detailed cost information is provided in **Supplementary Table S5**. Costs were adjusted to 2024 ZAR, where necessary, using the consumer price index, and then converted to United States Dollar (USD) at the January–July 2024 mean exchange rate (1 USD = 18.7 ZAR) [42,43]. All costs are presented in 2024 USD.

**Table 2:** Unit costs per patient screened or tested.

| Cost Category | USD (\$) |
| --- | --- |
| ***Syndromic management*** |  |
| STI symptom screen | 1.31 |
| Pelvic examination (clinical evaluation, females) | 3.36 |
| ***Treatment*** |  |
| UDS treatment | 0.74 |
| VDS treatment | 0.76 |
| NG treatment | 0.37 |
| CT treatment | 0.38 |
| ***Diagnostic testing*** |  |
| NG POCT (*TPP optimal*) | 9.11 |
| NG/CT POCT (*TPP minimal*) | 18.11 |
| NG/CT GeneXpert test | 48.98 |
| ***Visit costs*** |  |
| Primary STI visit (e.g., symptomatic) | 6.65 |
| Non-primary STI visit (e.g., asymptomatic) | 3.32 |
| Result delivery time | 1.31 |

### Cost-effectiveness analysis

Using the QALYs as the effect measure, we calculated the average cost-effectiveness ratios (ACERs) of the respective diagnostic strategies compared to SOC and the incremental cost-effectiveness ratios (ICERs) of the diagnostic strategies compared to each other. When analyzing the asymptomatic screening strategies, wherein there is no base case to compare, the cost-effectiveness ratio of the first least costly scenario was the total cost of that scenario divided by the QALYs gained in that scenario.

## Results

### Modeled STI incidence and prevalence

An estimated 7.6 million incident STI cases occurred in the modelled population aged 15-59 over the course of one-year, 4.1 million of which were among women and 3.5 million were among men. In 2017 there was an estimated 6.08 million STI cases among adults aged 15-49 in South Africa [2]. Of all infections, 3.58 million were symptomatic. Including all-cause incident cases of UDS and VDS, 3.15 million individuals sought care for STI-related symptoms in the public sector and were screened for NG/CT.

### Health outcomes of diagnostic strategies

Syndromic management, under the SOC, led to the least QALYs gained and cases treated, with the most cases missed and excess antibiotic use compared to diagnostic testing. The hypothetical NG/CT POCT had the best health outcomes with the least amount of excess antibiotic use and missed cases. The NG-only POCT had the second-best health outcomes, but also the second greatest amount of excess antibiotic use as NG-negative/CT-suspect cases were treated through syndromic management for CT which has poor specificity. GeneXpert testing for NG/CT had the least amount of excess antibiotic use but resulted in more missed cases, treating fewer cases than the POCTs, and gained the least number of QALYs due to LTFU for result delivery and treatment.

### Cost of diagnostic strategies

When considering the costs of care and treatment, syndromic management offers the lowest total cost and cost per case treated (**Table 3**). This was followed by the NG-only POCT and the NG/CT POCT, while Gene Xpert testing cost the most per case treated. Compared to syndromic management, the ACER per additional QALY gained were US$1,083 for the NG-only POCT,

**Table 3.** Health and economic outcomes by NG/CT diagnostic strategy for the entire national population of symptomatic cases in South Africa. The average cost-effectiveness ratio (ACER) is the additional cost per QALY gained beyond the SOC. The incremental cost-effectiveness ratio (ICER) is the cost per each additional QALY gained compared to the next least costly scenario. Percent (%) addition to budget is in addition to syndromic management under the SOC.

| Diagnostic Strategy | NG/CT Cases Treated | QALYs Gained | Excess Antibiotics | Total Cost (Million USD) | Cost/Case Treated (USD) | ACER/ QALY (USD) | ICER/ QALY (USD) | % Addition to the Budget |
| --- | --- | --- | --- | --- | --- | --- | --- | --- |
| SOC | 1,343,461 | 105,898 | 754,018 | \$ 34.1 M | \$ 25.40 | Ref. | Ref. | / |
| POCT NG | 1,599,376 | 126,948 | 391,805 | \$ 56.9 M | \$ 35.60 | \$1,083 | \$1,083 | 67% |
| POCT NG/CT | 1,673,939 | 133,302 | 118,539 | \$ 79.0 M | \$ 47.22 | \$1,638 | \$3,479 | 132% |
| Gene Xpert | 1,516,023 | 120,641 | 21,113 | \$ 198.5 M | \$ 130.97 | \$11,152 | Dominated | 482% |

US$1,638 for the NG/CT POCT, and US$11,152 for Gene Xpert. When comparing the diagnostic strategies incrementally, both the NG-only POCT and the NG/CT POCT were on the cost-effectiveness frontier and dominated GeneXpert testing. All the testing scenarios require a relatively substantial increase to the overall budget (in this case, the SOC): POCT scenarios require a 67-132% increase to the budget, with GeneXpert the most expensive, requiring a 5-fold increase to the budget.

### Sex-specific health outcomes

When results are stratified by sex, diagnostic testing had a greater impact in women compared to men due to the higher incidence in women (**Table 4**). Among men, the NG-only POCT treated 45,371 more cases and gained 3,176 more QALYs than the SOC. Among women, the NG-only POCT treated 210,543 more cases and gained 17,878 more QALYs. These differences were even greater for NG/CT POCT. GeneXpert also performed better than the SOC among women, treating 226,697 more cases and gaining 18,532 more QALYs. The cost per case treated using the NG-only or NG/CT POCTs was less than the SOC among women, wherein the costs of these diagnostic strategies were higher than the SOC in men. Both the ACERs and ICERs per additional QALY gained were significantly lower among women compared to men.

**Table 4.** NG/CT cases treated, QALYs gained, excess antibiotic use, cost per case treated, the average and incremental cost-effectiveness ratios for each additional QALY gained beyond the SOC (ACER, ICER), by diagnostic strategy when stratified by sex.

| Scenario | NG/CT Cases Treated | QALYs Gained | Excess Antibiotics | Cost/Case Treated (USD) | Total Cost (USD) | ACER/ QALY (USD) | ICER/ QALY (USD) |
| --- | --- | --- | --- | --- | --- | --- | --- |
| **Men** |  |  |  |  |  |  |  |
| SOC | 1,102,242 | 77,157 | 75,322 | \$ 10.38 | \$11.4 M | Ref. | Ref. |
| POCT NG | 1,147,613 | 80,333 | 48,837 | \$18.35 | \$21.1 M | \$ 3,026 | \$3,026 |
| POCT NG/CT | 1,157,655 | 81,036 | 32,514 | \$ 28.35 | \$32.8M | \$ 5,511 | \$16,741 |
| Gene Xpert | 1,048,107 | 73,367 | 6,138 | \$ 78.17 | \$81.9 M | Dominated | Dominated |
| **Women** |  |  |  |  |  |  |  |
| SOC | 241,219 | 28,741 | 678,697 | \$ 94.04 | \$22.7 M | Ref. | Ref. |
| POCT NG | 451,762 | 46,615 | 342,968 | \$79.42 | \$35.9 M | \$ 738 | \$738 |
| POCT NG/CT | 516,284 | 52,266 | 86,025 | \$ 89.52 | \$46.2 M | \$ 1,001 | \$1,830 |
| Gene Xpert | 467,916 | 47,273 | 14,975 | \$ 249.22 | \$116.6 M | \$ 5,069 | Dominated |

### Opportunistic screening scenarios of symptomatic & asymptomatic individuals

NG/CT screening could be implemented opportunistically in select use-cases using currently available GeneXpert testing or with the NG-only POCT once it becomes available. Under these scenarios, the opportunistic screening use-cases with the greatest potential impact were at ART clinics and during HIV testing. Combined, screening in these settings would result in the treatment of an estimated 50% of the annual NG burden with the NG-only POCT or 46% of the annual NG/CT burden with GeneXpert. These proportions would be even greater among women at 55% of the NG burden or 49% of the NG/CT burden. NG-only POCT screening in these settings would cost US$238 million annually or require a 697% increase in budget compared to SOC syndromic management, while GeneXpert would cost US$1.14 billion (3,333% budget increase; **Table 5**). The other possible settings for routine screening—family planning, PrEP, ANC—achieved considerably less coverage and proportion of total national NG/CT burden treated (**Table 5**). However, screening with the NG-only POCT during ANC could achieve 82% NG testing coverage and treat 79% of the NG burden among pregnant women, preventing 11,910 cases of preterm birth and vertical transmission of NG, gaining 4,790 QALYs, for US$7 million annually (20% budget increase; **Table 5**). GeneXpert screening during ANC could treat 87% of the NG/CT burden among pregnant women, preventing 43,045 cases of preterm birth and vertical transmission, gaining 12,902 QALYs, for US$33.5 million (**Table 5**).

**Table 5.** Cost-effectiveness analysis and incremental cost-effectiveness ratios (ICERs) of the opportunistic screening use-cases when NG-only sensitivity and specificity are 96.1% and 97.2% respectively. Dominated: health outcomes worse than previous strategy; weakly dominated: dominated by a more cost-effective strategy.

| Strategy | Test | Reduction in VT & PTB | QALYs | Total Cost (Million USD) | % addition to budget | ICER per QALY |
| --- | --- | --- | --- | --- | --- | --- |
| ANC | POCT NG | 11910 | 4943 | \$ 7,009,141 | 21% | \$1,417.88 |
| PrEP | POCT NG | 743 | 3759 | \$ 7,481,492 | 22% | Dominated |
| Family Planning | POCT NG | 0 | 20827 | \$ 39,430,080 | 116% | Weakly dominated |
| HIV Clinics | POCT NG | 2956 | 26663 | \$ 60,537,380 | 177% | Weakly dominated |
| HIV Testing | POCT NG | 9616 | 67880 | \$ 177,481,504 | 520% | Weakly dominated |
| ANC & Family Planning | POCT NG | 11910 | 25771 | \$ 46,439,221 | 136% | \$1,893.19 |
| HIV Clinics & Testing | POCT NG | 12572 | 94543 | \$ 238,018,884 | 697% | \$2,785.69 |
| ANC | GeneXpert NGCT | 43045 | 12902 | \$ 33,484,737 | 98% | Weakly dominated |
| PrEP | GeneXpert NGCT | 2684 | 8208 | \$ 35,740,321 | 105% | Dominated |
| Family Planning | GeneXpert NGCT | 0 | 46566 | \$ 188,374,810 | 552% | Dominated |
| **HIV Clinics** | GeneXpert<br>NGCT | 10682 | 59487 | \$<br>289,266,604 | 848% | Dominated |
| **HIV Testing** | GeneXpert<br>NGCT | 34756 | 152620 | \$<br>848,226,395 | 2485% | Weakly<br>dominated |
| **ANC &**<br>**Family**<br>**Planning** | GeneXpert<br>NGCT | 43045 | 59468 | \$<br>221,859,547 | 650% | Dominated |
| **HIV Clinics**<br>**& Testing** | GeneXpert<br>NGCT | 45438 | 212107 | \$<br>1,137,492,999 | 3333% | \$7,650.95 |

The cost-effectiveness analysis comparing NG-only and GeneXpert opportunistic screening use-cases and their ICERs per additional QALY gained are shown in **Table 5/Figure 1**. ANC, ANC and family planning, HIV clinics and testing, were all found to be on the cost-effectiveness frontier, in addition to HIV clinics and testing with GeneXpert (**Figure 1**). All other use cases were not found to be cost-effective for either the NG POCT or GeneXpert testing.

**Figure 1.**
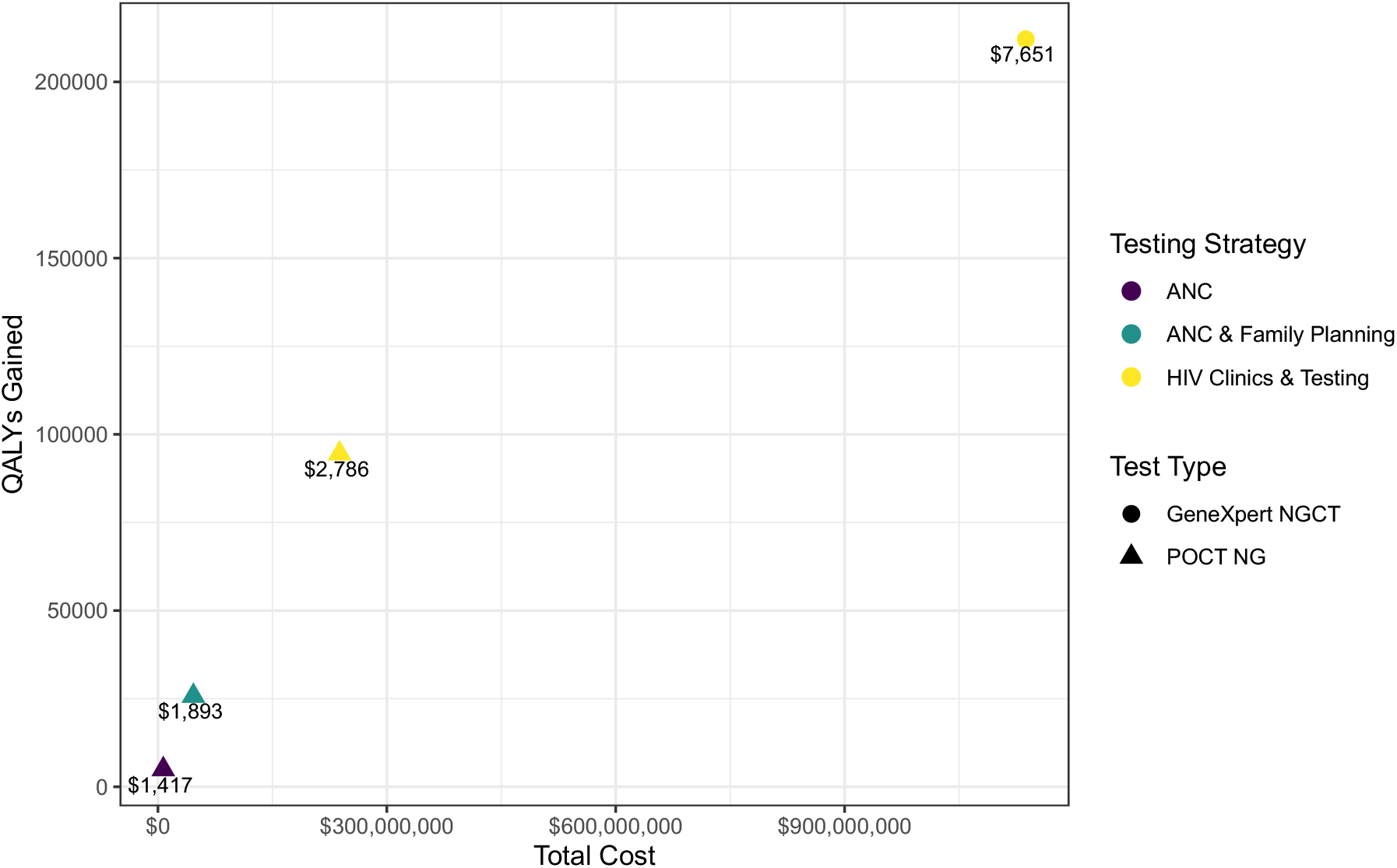
Opportunistic screening use-cases found to be on the cost-effectiveness frontier when comparing the NG-only POCT to GeneXpert testing. The corresponding labels are the incremental cost-effectiveness ratios. The reference scenario for this cost-effectiveness analysis was no asymptomatic screening.

### Sensitivity Analysis

Results from the sensitivity analyses are reported in **Text S2**.

## Discussion

Syndromic management under the SOC has relatively poor sensitivity and specificity— particularly among women—and leads to excessive antibiotic use [6,44]. Excess antibiotic use is especially problematic for *Neisseria gonorrhoeae* as ceftriaxone is currently the last line of empiric treatment, and resistance to this antibiotic is increasing. Zoliflodacin and gepotidacin are new treatments in the pipeline for NG, but initially may only be used in cases of treatment failure and/or drug-resistance, which makes preservation of existing and future antibiotics for gonorrhea crucial [10,45]. Therefore, there is an urgent need to understand the impact and costs of new STI diagnostic strategies in settings where syndromic management is the SOC, such as in South Africa, to inform policy and implementation strategies. This analysis shows that a potential POCT for NG or NG/CT would be effective and potentially cost-effective for symptomatic testing of STIs in South Africa, depending on the willingness to pay threshold per QALY gained.

Pichon-Riviere et al. estimated that the cost-effectiveness threshold (CE threshold) per QALY gained in South Africa was US$4,512 (3,292–7,941) in 2019 [46]. Our cost-effectiveness analysis showed that the incremental cost-effectiveness ratios per QALY gained for an NG-only POCT (US$1,083) or an NG/CT POCT (US$3,497) would be considered cost-effective under this threshold (approximately US$5,418 in 2024). Even under reduced performance (80% sensitivity and 95% specificity, as outlined in the minimum WHO TPP requirements [36]) the NG-only POCT would still be cost-effective under this metric with an ICER of US$2,723, dominating the NG/CT POCT. While the hypothetical POCT scenarios generally dominated GeneXpert testing in the analysis, these POCTs are not yet available. Given South Africa’s strong existing capacity for GeneXpert testing, it is reasonable to instead consider the ACER of GeneXpert. In this context, GeneXpert testing could be considered cost-effective for symptomatic women with an ACER of US$5,073 compared to the SOC. Although South Africa does not currently use an explicit cost-effectiveness threshold in decision-making, the threshold proposed by Pichon-Riviere et al. provides useful context for interpreting these results. However, despite favorable ICERs, all testing strategies represent a significant increase in total budget, with estimated increases ranging from 67% to 482% (US$23–164 M) compared to syndromic management, calling into question whether they would be considered affordable.

Other studies in the literature confirm the effectiveness and feasibility of etiological testing using GeneXpert for NG/CT among symptomatic cases in South Africa, but also the significantly increased cost. Lekodeba et al. found a similar cost per case treated (US$183) using near-POC GeneXpert for NG/CT symptomatic testing among adults in South Africa [47]. Marcus et al. found a similar cost per person tested using on-site GeneXpert testing (US$55.70) among youth seeking STI care in South Africa, but a much lower cost for lab-based testing (US$28.84) [35]. Smith et al. evaluated a new POC test for female genital inflammation, finding it to be cost-effective compared to syndromic management (ICER per case treated: US$11.08), even when considering a slightly higher syndromic management sensitivity (58–66%) [48]. Additionally, Smith et al. estimated a similar per patient cost for syndromic management (US$13.84) and a POCT cost (US$17.32) comparable to that of our NG POCT cost (US$15.76–19.12). Together with prior research from the literature, our results indicate a POCT for NG or NG/CT could be a cost-effective alternative to syndromic management that can greatly limit excess antibiotic use.

We further considered both an NG-only POCT and GeneXpert for opportunistic screening use-cases (given that they are available or nearly available)—implemented during ANC, PrEP initiation, family planning (contraceptive appointments), HIV testing, and at HIV clinics during ART follow-up. Our cost-effectiveness analysis showed that implementation of an NG-only POCT at ANC, ANC and family planning, and HIV clinics and testing would be cost-effective under the CE threshold. (**Figure 1**). The NG POCT screening scenarios remained on the cost-effectiveness frontier even when the sensitivity of the POCT was drastically reduced (65.8%, below the TPP requirement) to the level seen for the NG LFA among a population of asymptomatic pregnant women in Harare, Zimbabwe [49]. GeneXpert testing was only on the cost-effectiveness frontier for screening at HIV clinics and testing, but with nearly five times the budget required of the next least costly scenario on the cost-effectiveness frontier. The ICER ($7,651) was still under the upper-bound of the CE threshold.

An important limitation of this analysis is the inability to directly quantify the long-term health and economic consequences of excess antibiotic use. We must also acknowledge that expanded asymptomatic screening will ultimately lead to more unnecessary antibiotic use, in addition to its health benefits. While our model captures the number of unnecessary antibiotic courses administered under each scenario, it does not assign a cost or health penalty to this overuse— despite strong evidence that it contributes to AMR, with substantial implications for future treatment effectiveness and healthcare costs. The WHO has advocated for the use of an “antibiotic tax” to reflect these externalities, but estimating an appropriate value remains methodologically challenging [50]. We estimated the threshold antibiotic cost per course that would be required to make testing strategies cost-neutral compared to SOC. An NG/CT POCT would require a tax of approximately US$71 per excess antibiotic course to be cost-neutral, while GeneXpert would require a tax of US$224. POCT strategies—particularly the NG/CT LFA—achieve the greatest QALY gains but also lead to more excess antibiotic use than GeneXpert. Conversely, GeneXpert has the lowest excess antibiotic use due to its high specificity, and if the societal costs of AMR could be more accurately quantified and included, GeneXpert would likely be viewed more favorably in cost-effectiveness terms, despite its limitations. Until these downstream consequences are incorporated, cost-effectiveness analyses may undervalue the full public health benefit of diagnostic strategies that reduce antibiotic overuse.

This study comes with some additional limitations. For one, in some cases (e.g., incidence) we were limited by data that was publicly available and had to make broad population level estimates—that were not specific to key populations. The performance of syndromic management flowcharts under SOC for NG/CT detection is likely to vary across settings and we cannot be certain of the sensitivity and specificity in South Africa, so we provide a sensitivity analysis around these estimates. Similarly, until further studies are done on the NG LFA, we will not know the true performance, but we provide a sensitivity analysis around estimates seen in a symptomatic population, an asymptomatic population, and those outlined in the WHO TPP. Additionally, we assumed the same accuracy for the NG-only and NG/CT POCTs, but in practice an NG/CT POCT may have different performance. The opportunistic screening use-cases we present are not additive to each other or to symptomatic testing, as the overlap between the populations seeking care through these avenues is unknown. This limited our ability to perform a cost-effectiveness analysis between use-cases and the SOC. We did however consider the combinations of use-cases which would not overlap—ANC and family planning or HIV clinics and testing. Furthermore, we assumed population prevalence among the populations seeking care through the use-cases as we expect the difference between prevalence and incidence to represent the cases caught through symptomatic care-seeking. Finally, for the cost of the POCT, we used the costs outlined in the WHO TPP, as this test has not yet come to market. As with all modelling studies, the results are context specific based on the chosen parameters, but this modelling framework could easily be modified to the population proportions and NG/CT epidemiology of other countries.

Future research would be required to understand the long-term impact of NG/CT screening strategies on NG/CT transmission, but this analysis shows the immediate benefit of a potential POCT with respect to infections treated, QALYs gained, and excess antibiotic use reduced. While this POCT is not yet available and its performance unknown, GeneXpert testing for NG/CT could be considered in some scenarios, such as among symptomatic women or during ANC to prevent pregnancy-related complications.

While syndromic management is low cost, it falls short in accuracy and contributes to excess antibiotic use. Diagnostic testing—though requiring greater investment—has a critical role to play in improving treatment outcomes and advancing antibiotic stewardship. As South Africa prepares for the introduction of novel antibiotics for gonorrhoeae such as zoliflodacin and gepotidacin, strengthening diagnostic capacity is essential to preserve antimicrobial effectiveness and ensure more targeted, cost-effective care, particularly for women.

## Data availability

The models and data used in this study are publicly available at https://doi.org/10.6084/m9.figshare.c.8082232.v1. All data and references used in the parameterization of the models are contained within this manuscript and the supplementary appendix.

