## Supplementary Appendix for "National scale-up of etiological testing for *N. gonorrhoeae* and *C. trachomatis* in South Africa: a health economic modelling analysis"

**Table S1.** Consolidated Health Economic Evaluation Reporting Standards (CHEERS) checklist.

| Section/topic | Item No | Guidance for reporting | Reported in section |
|---|---|---|---|
| **Title** | | | |
| Title | 1 | Identify the study as an economic evaluation and specify the interventions being compared. | Title |
| **Abstract** | | | |
| Abstract | 2 | Provide a structured summary that highlights context, key methods, results, and alternative analyses. | Abstract |
| **Introduction** | | | |
| Background and objectives | 3 | Give the context for the study, the study question, and its practical relevance for decision making in policy or practice. | Introduction |
| **Methods** | | | |
| Health economic analysis plan | 4 | Indicate whether a health economic analysis plan was developed and where available. | Methods |
| Study population | 5 | Describe characteristics of the study population (such as age range, demographics, socioeconomic, or clinical characteristics). | Methods |
| Setting and location | 6 | Provide relevant contextual information that may influence findings. | Introduction/ Methods/ Discussion |
| Comparators | 7 | Describe the interventions or strategies being compared and why chosen. | Introduction/ Methods |
| Perspective | 8 | State the perspective(s) adopted by the study and why chosen. | Methods |
| Time horizon | 9 | State the time horizon for the study and why appropriate. | Methods |
| Discount rate | 10 | Report the discount rate(s) and reason chosen. | Methods |
| Selection of outcomes | 11 | Describe what outcomes were used as the measure(s) of benefit(s) and harm(s). | Methods |
| Measurement of outcomes | 12 | Describe how outcomes used to capture benefit(s) and harm(s) were measured. | Methods |

| Section/topic | Item No | Guidance for reporting | Reported in section |
|---|---|---|---|
| Valuation of outcomes | 13 | Describe the population and methods used to measure and value outcomes. | Methods |
| Measurement and valuation of resources and costs | 14 | Describe how costs were valued. | Methods/ Supplement |
| Currency, price date, and conversion | 15 | Report the dates of the estimated resource quantities and unit costs, plus the currency and year of conversion. | Methods/ Supplement |
| Rationale and description of model | 16 | If modelling is used, describe in detail and why used. Report if the model is publicly available and where it can be accessed. | Methods/ Data availability |
| Analytics and assumptions | 17 | Describe any methods for analysing or statistically transforming data, any extrapolation methods, and approaches for validating any model used. | Methods |
| Characterising heterogeneity | 18 | Describe any methods used for estimating how the results of the study vary for subgroups. | Methods/ Discussion/ Supplement |
| Characterising distributional effects | 19 | Describe how impacts are distributed across different individuals or adjustments made to reflect priority populations. | Methods/ Results |
| Characterising uncertainty | 20 | Describe methods to characterise any sources of uncertainty in the analysis. | Methods/ Results/ Supplement |
| Approach to engagement with patients and others affected by the study | 21 | Describe any approaches to engage patients or service recipients, the general public, communities, or stakeholders (such as clinicians or payers) in the design of the study. | N/A |
| **Results** | | | |
| Study parameters | 22 | Report all analytic inputs (such as values, ranges, references) including uncertainty or distributional assumptions. | Methods/ Supplement |
| Summary of main results | 23 | Report the mean values for the main categories of costs and outcomes of interest and summarise them in the most appropriate overall measure. | Results |
| Effect of uncertainty | 24 | Describe how uncertainty about analytic judgments, inputs, or projections affect | Results/ Discussion/ |

| Section/topic | Item No | Guidance for reporting | Reported in section |
|---|---|---|---|
| | | findings. Report the effect of choice of discount rate and time horizon, if applicable. | Supplement |
| Effect of engagement with patients and others affected by the study | 25 | Report on any difference patient/service recipient, general public, community, or stakeholder involvement made to the approach or findings of the study | N/A |
| **Discussion** | | | |
| Study findings, limitations, generalizability, and current knowledge | 26 | Report key findings, limitations, ethical or equity considerations not captured, and how these could affect patients, policy, or practice. | Discussion |
| **Other relevant information** | | | |
| Source of funding | 27 | Describe how the study was funded and any role of the funder in the identification, design, conduct, and reporting of the analysis | Funding |
| Conflicts of interest | 28 | Report authors conflicts of interest according to journal or International Committee of Medical Journal Editors requirements. | Conflicts of Interest |

### Text S1. Supplementary Methods

*Sensitivity analyses*

We performed several sensitivity analyses around key parameters in this analysis. The first considers South African guidelines with respect to STI-treatment, in which all individuals presenting with UDS and VDS receive empiric treatment for all possible etiologies, regardless of clinical evaluation or judgement (i.e., all patients with UDS/VDS receive NG/CT treatment). The second considers a SOC scenario where the sensitivity of syndromic management for VDS was increased to 83.9% and specificity decreased to 45.3% as observed in Wi et al. [1]. In a sensitivity analysis for the LFA, we reduced the sensitivity and specificity of symptomatic testing to 80% and 95%, respectively, based on the minimum criteria outlined in the WHO TPP for a rapid NG POCT [2]. While for the asymptomatic screening scenarios, we reduced the sensitivity to 65.8% and increased the specificity to 99.2%, based on a screening study among predominantly asymptomatic pregnant women attending ANC in Harare, Zimbabwe [3]. We evaluated the impact of the rate of GeneXpert-related LTFU on health outcomes, reducing LTFU to 0%. As an additional sensitivity analysis, we examined the cost threshold at which reducing excess antibiotic use would render etiological testing strategies cost-neutral compared to syndromic management. Specifically, we estimated the required per-dose "antibiotic tax"—a hypothetical monetary value representing the long-term health and societal costs of unnecessary antibiotic use—that would equalize the ACERs of testing strategies with the SOC.

**Table S2**. Opportunistic use-case care-seeking behavior by key population.

| **Population** | **HIV Testing** | **HIV Care (ART)** | **PrEP** | **Family Planning** | **ANC** |
|---|---|---|---|---|---|
| AGYW | 52.5% [4] | 51.0% [5] | 6.66% [5] | 22.4% [4] | - |
| -Women >25 | 58.5% [4] | 78.0% [5] | 0.87% [5] | 22.7% [4] | - |
| Pregnant | 94.5% [4] | 98.0% [6] | 6.50% [7] | - | 94% [4] |
| FSW | 58.5% [4] | 73.0% [5] | 49.37% [5] | Could FSW access FP services? | - |
| MSW | 44.1% [4] | 70.0% [5] | 0.63% [5] | - | - |
| MSM/TGW | 48.0% [8] | 62.0% [5] | 22.10%, 11% [5,9] | - | - |
| Public sector care-seeking | 91% [10] | 91% [10] | 100% | 74% [4] | 87% [4] |

**Table S3**. Risk of pregnancy and birth related complications associated with NG or CT infection. PID: pelvic inflammatory disease.

| **Parameter** | **Risk if untreated** |
|---|---|
| PID, NG | 20% [11] |
| PID, CT | 10% [12] |
| PID, baseline | 3.1% [13] |
| Vertical transmission, NG | 29% [14] |
| Vertical transmission, CT | 43% [14] |
| Pre-term birth, NG | 12.2% [15] |
| Pre-term birth, CT | 9.9% [15] |
| Pre-term birth, baseline | 7.9% [15] |

**Table S4**. Utility scores for the quality adjusted life-years associated with NG/CT related health states. PID: pelvic inflammatory disease.

| **Test** | **Utility score** |
|---|---|
| Asymptomatic infection | 1.00 [16] |
| Symptomatic infection | 0.93 [16] |
| PID | 0.80 [16] |
| Vertical Transmission | 0.904 [17,18] |
| Pre-term birth | 0.94 [19,20] |

### Table S5: Cost assumptions

| Unit | Cost (USD) | Assumption | Source |
|---|---|---|---|
| STI symptom screen | $1.31 | 5 min of a professional nurse's time to relay test results, provide treatment if necessary | https://www.dpsa.gov.za/policy-updates/nlrrm/remuneration_policy/annual_cost_of_living_adjustments/ https://www.medicalinnovations.co.za/product/vaginal-speculum/ |
| Pelvic examination | $3.36 | 10 min of a professional nurse's time to perform examination; speculum | |
| Result delivery | $1.31 | 5 min of a professional nurse's time to relay test results, provide treatment if necessary | https://www.dpsa.gov.za/policy-updates/nlrrm/remuneration_policy/annual_cost_of_living_adjustments/ |
| Primary STI visit | $6.65 | Overheads, assets, non-clinical staff costs per patient visit | Cost and outcomes of routine HIV care and treatment: public and private service delivery models covering low-income earners in South Africa - PMC (nih.gov). https://pubmed.ncbi.nlm.nih.gov/36906559/ |
| Non-primary STI visit | $3.32 | Overheads, assets, non-clinical staff costs (assumed 50% of time allocated to STI) per patient visit | Cost and outcomes of routine HIV care and treatment: public and private service delivery models covering low-income earners in South Africa - PMC (nih.gov) https://pubmed.ncbi.nlm.nih.gov/36906559/ |
| UDS treatment | $0.74 | 1g ceftriaxone, 100mg doxycycline (14 tablets) | WHO Guidelines (July 2024) https://www.who.int/publications/i/item/9789240090767 , Master Health Product List (Feb 2024) Tenders – National Department of Health |
| VDS treatment | $0.76 | 1g ceftriaxone, 100mg doxycycline (14 tablets), 2g metronidazole | |
| NG treatment | $0.37 | 1g ceftriaxone | WHO Guidelines (July 2024) https://www.who.int/publications/i/item/9789240090767 , Master Health Product List (Feb 2024) Tenders – National Department of Health |
| CT treatment | $0.38 | 100mg doxycycline | |
| NG POCT | $9.11 | 15 min of a professional nurse's time to collect sample, conduct test, provide result; + $3.00 USD POCT; + training (90 min test training + 1 day guideline training) | https://www.dpsa.gov.za/policy-updates/nlrrm/remuneration_policy/annual_cost_of_living_adjustments/, WHO Target product profile for POC tests for STIs, www.finddx.org |
| NG/CT POCT | $18.11 | 15 min of a professional nurse's time to collect sample, conduct test, provide result; + $12.00 USD POCT; + training (90 min test training + 1 day guideline training) | |
| NG/CT GeneXpert test | $48.98 | GeneXpert MTB/XDR implementation in South Africa (including cost of consumables, equipment, registration, transportation, staff, and training) - MTB/XDR cartridge + NG/CT cartridge + 10% overheads + 3.69% EQA + 3% error rate. Including 5 min of a professional nurse's time to collect sample and training on | Cassim et al., 2024, https://doi.org/10.5588/ijtldopen.23.0501, https://dxc-marketplace.finddx.org/pages/cepheid-accessible-pricing |

| | | |
|---|---|---|
| | | etiological management (1 day of training for 50% of professional nurses in South Africa, valid for 5 years) |

### Text S2. Supplementary Results

*Sensitivity analysis*

A POCT with specifications reduced to the minimal TPP requirements (80% sensitivity, 95% specificity) still treats more cases, gains more QALYs, and reduces more excess antibiotic use than the SOC for both the NG-only or NG/CT POCT (**Table S6**). However, the improved health outcomes only extend to women, as the reduced sensitivity leads to fewer cases treated and QALYs gained among men compared to SOC. Therefore, the incremental cost per additional QALY gained over SOC became dominated for men, while increasing to US$1,109 for the NG-only POCT. The NG/CT POCT became dominated compared to GeneXpert testing overall and among men, while increasing to US$2,566 among women.

If SOC performance was modified for VDS, effectively increasing the sensitivity to 83.9% and decreasing the specificity to 45.3%, more cases are treated and QALYs are gained, but excess antibiotic use increased compared to SOC at baseline. While both the NG-only and NG/CT POCT would still have better health outcomes and less antibiotic use than SOC, the NG-only POCT would become dominated by the NG/CT POCT. If the POCT sensitivity was reduced to 80% as in the minimal TPP requirements, it would be dominated by SOC.

If we assume complete empiric treatment under the SOC—meaning all symptomatic cases receive treatment for both gonorrhea and chlamydia—it would cost the least and effectively treat all symptomatic cases of STIs seeking care. However, it would also lead to extreme excess antibiotic use—approximately 2.8 million doses of excess and unnecessary antibiotics would be administered, half of which would be ceftriaxone.

If we assume there is no LTFU between GeneXpert testing and result delivery, both health outcomes and cost efficiencies improve for the GeneXpert testing scenario (**Table S7**), although the total cost of the strategy increases as more cases return for result delivery. The health outcomes are comparable, but the number of QALYs gained remain less than with NG/CT POCT, meaning that GeneXpert remains dominated in the cost-effectiveness analysis. When considering a reduced sensitivity and specificity of the NG POCT in largely asymptomatic populations, such as in the opportunistic screening scenarios, the same scenarios remain on the cost-effectiveness frontier (**Figure S4**).

Finally, we estimated the minimum antibiotic tax per antibiotic course prescribed required for each testing strategy to be cost-neutral compared to syndromic management. This value reflects the breakeven point at which the societal cost of unnecessary antibiotic use justifies investment in diagnostic testing, independent of QALY gains. For the NG-only POCT, a per-course tax of US$63 would be needed, for NG/CT POCT US$71 would be needed, while for GeneXpert testing, the required tax would be US$224 per excess antibiotic course avoided (**Table S8**).

**Table S6.** Sensitivity analysis when the POCT sensitivity and specificity are reduced to TPP values of 80% and 95%, respectively.

| Diagnostic Strategy | NG/CT Cases Treated | QALYs Gained | Excess Antibiotics | Total Cost (Million USD) | Cost/Case Treated (USD) | ACER/ QALY (USD) | ICER/ QALY (USD) |
|---|---|---|---|---|---|---|---|
| SOC | 1,343,461 | 105,818 | 754,018 | $ 34.1 M | $ 25.40 | Ref. | Ref. |
| POCT NG | 1,445,262 | 114,288 | 403,431 | $ 57.2 M | $ 39.57 | $2,723 | $2,723 |
| POCT NG/CT | 1,393,498 | 112,228 | 211,677 | $ 79.8 M | $ 57.28 | $7,127 | Dominated |
| Gene Xpert | 1,516,023 | 120,543 | 21,113 | $ 198.5 M | $ 130.97 | $11,260 | $22,599 |

**Table S7.** Sensitivity analysis when GeneXpert LTFU is reduced to 0%.

| Diagnostic Strategy | NG/CT Cases Treated | QALYs Gained | Excess Antibiotics | Total Cost (Million USD) | Cost/Case Treated (USD) | ACER/ QALY (USD) | ICER/ QALY (USD) |
|---|---|---|---|---|---|---|---|
| SOC | 1,343,461 | 105,818 | 754,018 | $ 34.1 M | $ 25.40 | Ref. | Ref. |
| POCT NG | 1,599,376 | 126,872 | 391,805 | $ 56.9 M | $ 35.60 | $1,083 | $1,083 |
| POCT NG/CT | 1,673,939 | 133,193 | 118,539 | $ 79.0 M | $ 47.22 | $1,691 | $3,497 |
| Gene Xpert | 1,647,851 | 131,025 | 22,949 | $ 200.6 M | $ 121.70 | $6,602 | Dominated |

**Table S8:** Estimated antibiotic penalty required for etiological testing strategies to achieve cost neutrality with syndromic management. The "required antibiotic penalty" represents the per-course cost of excess antibiotics that would need to be assigned to equalize the total cost of each testing strategy with syndromic management (SOC). This value reflects the breakeven point at which the societal cost of unnecessary antibiotic use justifies investment in diagnostic testing, independent of QALY gains.

| Scenario | Excess Antibiotics | Total Cost (Million USD) | Required Antibiotic penalty (USD) |
|---|---|---|---|
| SOC | 754,018 | $ 34.1 M | / |
| POCT NG | 391,805 | $ 56.9 M | $62.96 |
| POCT NGCT | 118,539 | $ 79.0 M | $70.67 |
| Xpert NGCT | 21,113 | $ 198.5 M | $224.34 |

**Figure S1**. Total *Neisseria gonorrhoeae* burden (grey) and burden treated (color) by target population, when an NG-only POCT is implemented at ART clinics (brown) and during HIV testing (green). Populations are inclusive of overlap between groups.

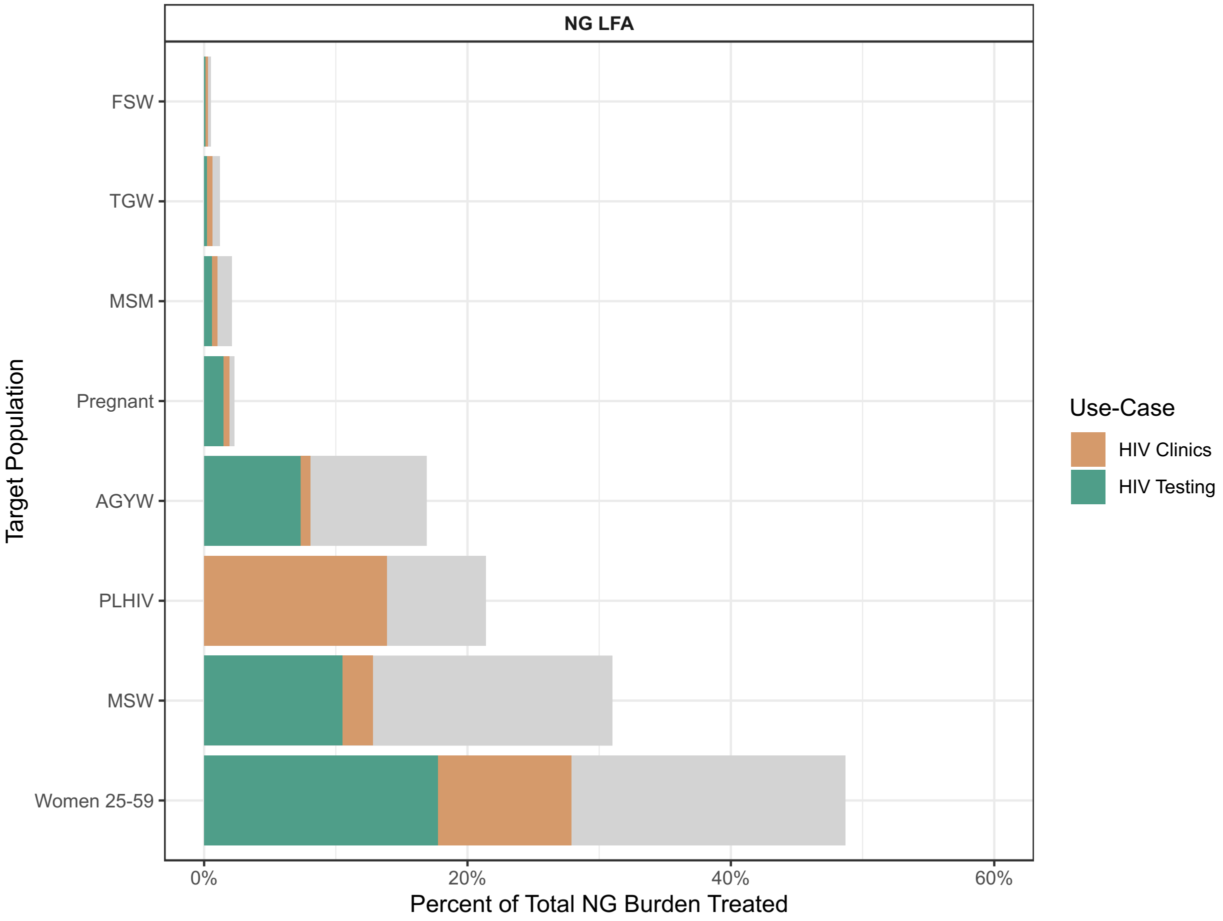

**Figure S2**. Total *Neisseria gonorrhoeae* burden (grey) and burden treated (color) by target population, when an NG-only POCT is implemented at family planning clinics (pink) and during antenatal care (purple). Populations are inclusive of overlap between groups.

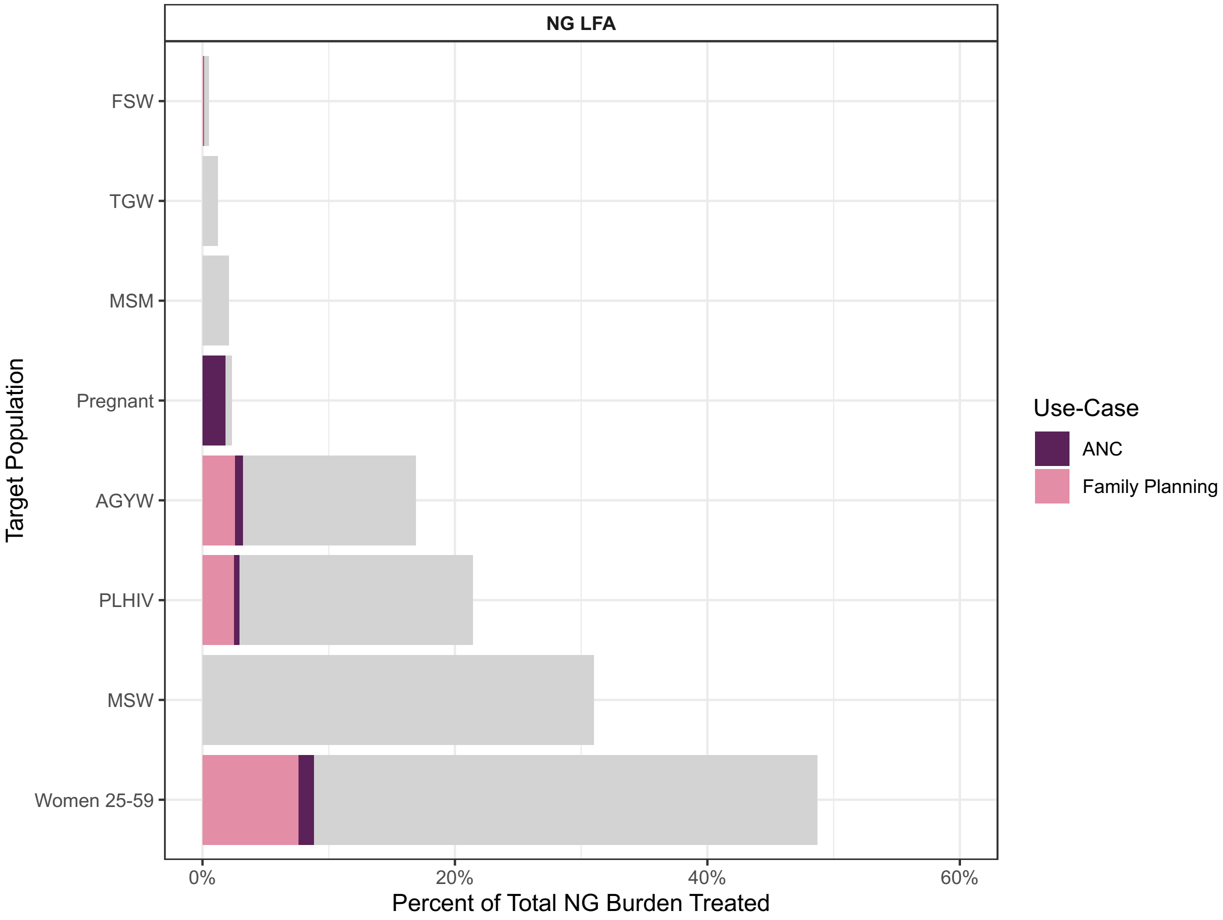

**Figure S3**. Total *Neisseria gonorrhoeae* burden treated (blue) by target population, when an NG-only POCT is implemented during pre-exposure prophylaxis (PrEP) initiation appointments. Populations are inclusive of overlap between groups.

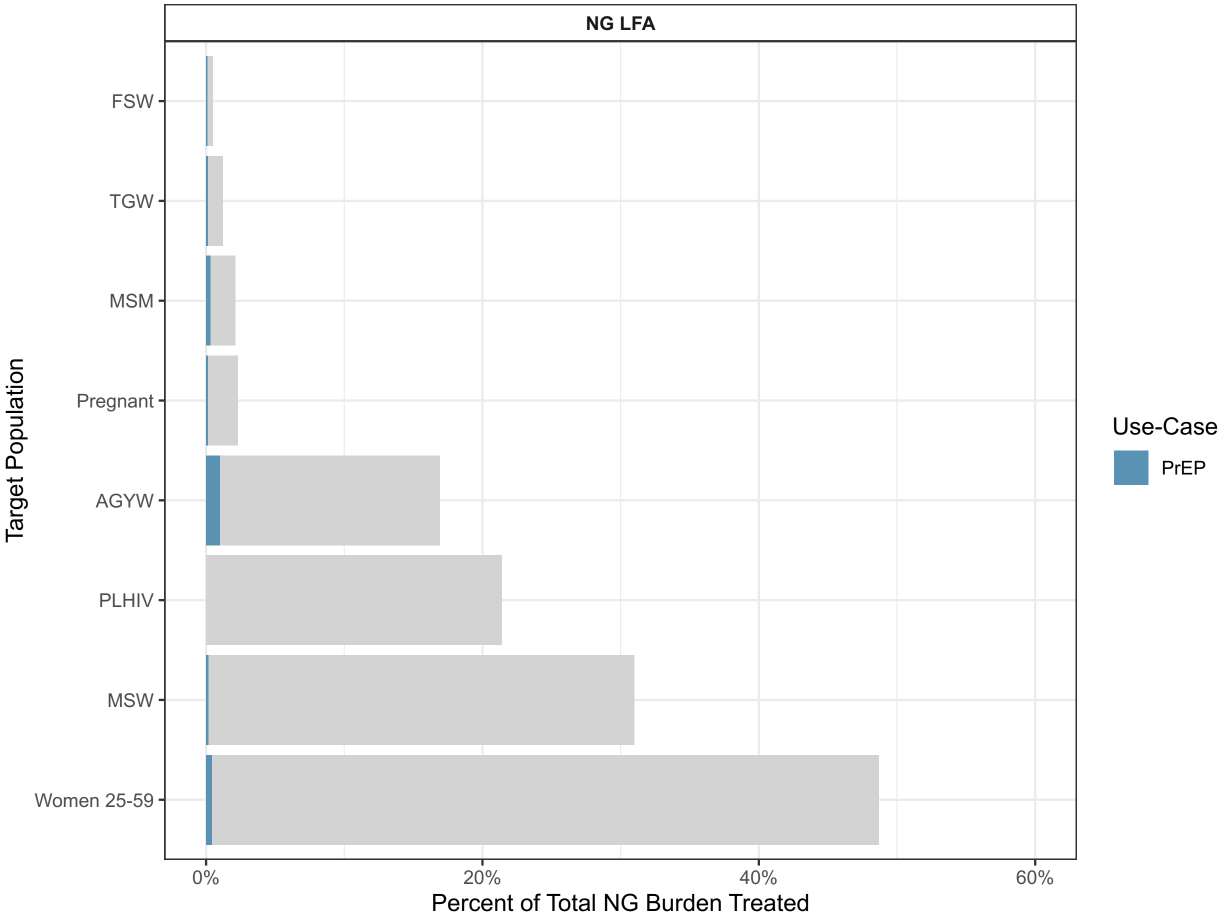

**Figure S4**. Opportunistic screening use-cases found to be on the cost-effectiveness frontier when comparing the NG-only POCT to GeneXpert testing. The NG-only POCT specifications have been reduced to 65.8% sensitivity and 99.2% specificity. The corresponding labels are the incremental cost-effectiveness ratios. The reference scenario for this cost-effectiveness analysis was no asymptomatic screening.

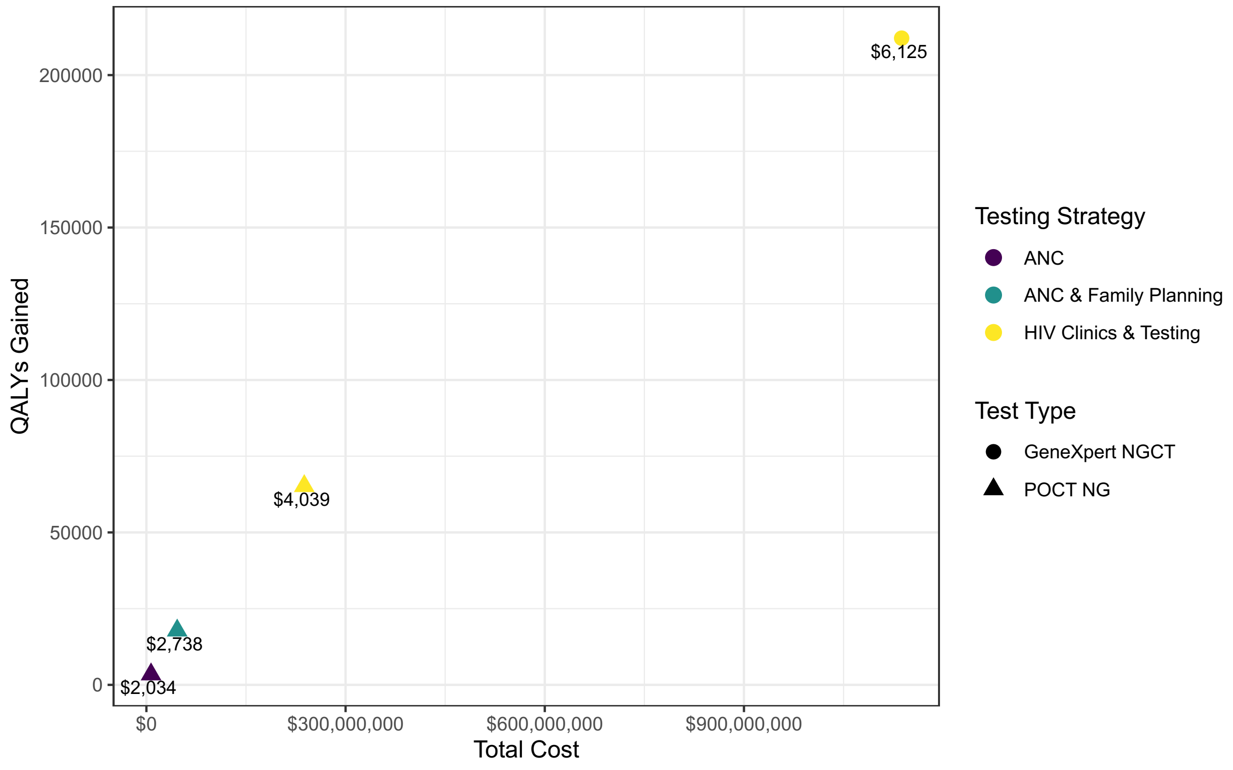
